# The Evolution of Long COVID Research, 2020 to 2025: A Bibliometric Analysis with Implications for Clinical Practice and Policy

**DOI:** 10.1101/2025.09.23.25336485

**Authors:** Lanre Peter Daodu, Noah Ajanaku, Kingsley Ikhuosho Ojeikere, Godwin E. Ozokolie, Methodius Okouzi, Oluwagbenga John Ogunbiyi, Chinedum Michael Onah, Muriana Olotu, Henry Chukwuebuka Chibuoke, Mary Erameh

## Abstract

**Background:** Since 2020, recognition of long COVID (post-acute sequelae of COVID-19) has prompted rapid multidisciplinary research across medicine, public health, psychology, and the social sciences. The literature is extensive and heterogeneous, making it challenging to synthesise current knowledge, identify evidence gaps, and inform research priorities.

**Methods:** We performed a bibliometric review of long COVID publications indexed in Scopus from January 2020 to March 2025. Records were analysed using VOSviewer and Biblioshiny to assess publication trends, country and institutional contributions, authorship networks, citation impact, and thematic clusters.

**Results:** Publications on long COVID grew at an annual rate of 19.81%, peaking in 2024, reflecting intensified global attention. The United States, China, and the United Kingdom were the largest contributors. Thematic mapping revealed three dominant clusters: clinical research on manifestations and diagnostics; psychological research on mental health and cognitive sequelae; and social and rehabilitation research addressing disability, return to work, and service delivery. Prominent institutions and key authors played a central role in knowledge production and collaboration networks. Notable gaps included limited studies on paediatric long COVID, scarce long-term outcome data from diverse populations and settings, and few rigorous evaluations of interventions and service models.

**Conclusions:** The long COVID research landscape from 2020 to 2025 is dynamic and increasingly collaborative, yet exhibits geographic and topical imbalances. To advance understanding and improve patient outcomes, stakeholders should support inclusive international collaborations, prioritise funding for underexplored populations and intervention trials, and accelerate translation of findings into clinical guidelines and integrated care models. Continued bibliometric monitoring will help target research investments and policy responses.

## Background

Long COVID, also referred to as post-acute sequelae of COVID-19 (PASC), typically describes the continuation or onset of symptoms lasting three months or more after the acute phase of SARS-CoV-2 infection. On 11^th^ June 2024, the National Academies of Sciences, Engineering, and Medicine (NASEM) established a new consensus definition of long COVID as an infection-associated chronic condition (IACC) that occurs after SARS-CoV-2 infection and persists for at least three months, presenting as a continuous, relapsing and remitting, or progressive disease state affecting one or more organ systems (1). It is a multisystem condition with heterogeneous presentations and trajectories. Recent large-scale evidence synthesises a substantial global burden. An updated systematic review and meta-analysis of 429 studies estimated a pooled global prevalence of 36% after SARS-CoV-2 infection, with higher estimates in South America (51%) (2). The prevalence persisted with time, at 35% before one year and 46% at one to two years of follow-up, highlighting ongoing morbidity beyond the acute illness (2). A recent policy-focused scoping review highlights the rapid expansion of reviews on long COVID and the need for consistent definitions, standardised outcome measures, and globally representative cohorts (3). Persistent heterogeneity across populations and methods complicates inference, reinforcing priorities for longitudinal follow-up, phenotype-specific trials, and evaluation of care pathways that integrate rehabilitation, mental health, and social support (3).

Following the 2020 recognition of long COVID, research on this complex and multisystem disease has rapidly expanded across medicine, public health, psychology, and the social sciences. This research is vast and diverse, spanning numerous disciplines, making it challenging to gain a comprehensive understanding of what is known, what remains unknown, and where the field is headed. This highlights the necessity of a rigorous and systematic analysis of the extant scientific literature on long COVID to guide future research and policy.

Bibliometric analysis, a quantitative method for evaluating research output, impact, and trends, offers a powerful tool to address this need (4). Bibliometric analyses provide valuable insights into the growth and evolution of research fields by examining publication patterns, citation networks, and thematic trends (5). Some bibliometric reviews have been conducted on COVID-19 research broadly (6, 7), only a few have focused explicitly on long COVID research, particularly over a defined period that captures its emergence and rapid development.

Moreover, the rapid proliferation of long COVID research has led to a fragmented and heterogeneous body of literature, making it difficult for researchers, policymakers, and clinicians to identify key findings and prioritise research areas (8, 9). The interdisciplinary nature of long COVID research presents both opportunities and challenges. While it fosters collaboration and innovation, it also raises questions about how different disciplines contribute to the field and whether there are disparities in research output and impact across regions and institutions (10, 11).

The main objective was to evaluate the trends and growth of long COVID research from 2020 to March 2025; this provided a macro-level understanding of how the research on the debilitating disease has expanded since its emergence. We provided answers to the following research questions (RQs): What are the trends and growth patterns in long-term COVID research from 2020 to March 2025? - This question was answered by analysing the volume and trajectory of research publications over time, identifying periods of significant increase or decrease. It helped understand how the scientific community’s focus on the disease has evolved since the pandemic began. Another RQ: Who were the most influential authors, institutions, and journals in the field of long COVID research? This question highlighted the major players driving advancements in long COVID research and their contributions. Also, what were the predominant research topics, themes, and methodologies used in long COVID studies? This question categorised and analysed the main areas of focus within long COVID research, including common themes and methodological approaches. It also provided insights into the intellectual structure of the field and identified dominant research trends. Finally, how has long COVID research impacted the scientific community, as evidenced by citation analysis and co-authorship networks? This question examined the influence and dissemination of long COVID research by analysing citation patterns and co-authorship networks. It shed light on how knowledge about long COVID was shared and utilised within the scientific community, indicating the research’s visibility and impact.

This bibliometric review utilised an extensive bibliographic database, Scopus, which offered comprehensive coverage of scientific literature across various disciplines. The review’s results would not only enhance academic knowledge of long COVID but also provide real-world applications for researchers and public health initiatives. The dissemination of scientific knowledge regarding long COVID and how collaborations shape the field will be better understood through this study’s analysis of citation patterns alongside co-authorship networks (4, 12).

## Methods

This study employed a bibliometric review to systematically analyse the research landscape of long COVID from 2020 to 2025. The methodology integrated two tools, VOSviewer and Biblioshiny (from the Bibliometrix R package), to ensure a comprehensive and robust analysis. These tools complement each other, with VOSviewer excelling in visualisation and cluster analysis (13, 14), and Biblioshiny provides advanced data preprocessing, statistical analysis, and network mapping capabilities.

### Data collection

We extracted bibliographic data from Scopus databases. Scopus is widely recognised for its comprehensive coverage of high-quality, peer-reviewed literature across disciplines. (15). This database was selected because it provides robust metadata, including author affiliations, citations, keywords, and abstracts, which are essential for bibliometric analysis (16).

### Search strategy

A systematic search was conducted on 14^th^ March 2025, using a combination of keywords and Boolean operators to capture the breadth of long COVID research. The search query included terms such as “long COVID”, “post-COVID syndrome, “post-acute COVID-19”, “chronic COVID”, “long-haul COVID”, and “PASC” (post-acute sequelae of SARS-CoV-2 infection). The search strings for this study were designed to capture the breadth of research on long COVID, including its various terminologies and manifestations. The search strategy employs Boolean operators (AND, OR) and truncation symbols (*) to ensure comprehensive coverage of the literature.

The search strings for the Scopus database were “(“long COVID” OR “long-COVID” OR “long COVID-19” OR “long-haul COVID” OR “long-haul COVID-19” OR “post-COVID syndrome” OR “post-COVID-19 syndrome” OR “post-acute COVID” OR “post-acute COVID-19” OR “chronic COVID” OR “chronic COVID-19” OR “PASC” OR “post-acute sequelae of SARS-CoV-2” OR “post-COVID condition” OR “post-COVID-19 condition” OR “ongoing COVID” OR “living with COVID” OR “long-tail COVID”) AND (“symptoms” OR “fatigue” OR “brain fog” OR “respiratory symptoms” OR “cardiovascular symptoms” OR “neurological symptoms” OR “cognitive dysfunction” OR “mental health” OR “multisystem inflammation” OR “children” OR “adults” OR “elderly” OR “healthcare workers” OR “comorbidities” OR “immunocompromised” OR “vaccination” OR “reinfection” OR “clinical trials” OR “cohort studies” OR “longitudinal studies” OR “qualitative research” OR “systematic reviews” OR “meta-analysis” OR “bibliometric analysis”)”. The inclusion of diverse terminologies related to the disease ensures that the search captures the full scope of research on the topic. The search was limited to articles published between January 2020 and March 2025, providing coverage of the entire period since the emergence of the COVID-19 pandemic. Only articles and reviews in English were included, while editorials, letters, and non-scientific publications were excluded to maintain the quality and relevance of the dataset.

### Bibliometric analysis

We employed a comprehensive suite of bibliometric techniques to rigorously address the research objectives and research questions, leveraging the complementary strengths of VOSviewer and Biblioshiny. Biblioshiny was selected because it was instrumental in quantifying the annual publication output, calculating growth trajectories, and mapping the disciplinary and geographical distribution of research on long COVID. Biblioshiny provided robust statistical summaries and visual trend analyses, which were critical for capturing the temporal dynamics of the field. Key performance indicators, including publication counts, citation metrics, and annual growth rates, were computed to assess the scholarly impact and productivity of authors, institutions, and journals. In addition, Biblioshiny facilitated the examination of collaboration patterns, offering insights into the structure and extent of research partnerships.

For thematic and intellectual mapping, VOSviewer was used to conduct co-word analyses and keyword clustering, allowing for the identification of dominant research themes and emerging topics. Its advanced clustering algorithms and visualisation capabilities proved particularly effective in delineating the conceptual landscape of long COVID research. Additionally, VOSviewer was used to construct citation and co-authorship networks, visually representing the interconnections among researchers, institutions, and countries. These visualisations were further enriched by Biblioshiny’s statistical insights into citation behaviour and collaborative dynamics, resulting in a nuanced and multidimensional understanding of the research ecosystem.

## Results and Discussion

### Publication trends and growth analysis

Table 1 presents an overview of the long COVID research landscape from 2020 to March 2025, reflecting the field’s rapid emergence and continued evolution. A total of 9,729 documents were retrieved from Scopus, with 80.9% comprising original research articles (n = 7,867) and 19.1% review articles (n = 1,862). This distribution highlighted a strong emphasis on primary research, supported by a substantial body of secondary literature synthesising existing findings. The annual publication growth rate was calculated at 19.81%, indicating a sustained and accelerating scholarly interest in long COVID. The average document age of 2.13 years further highlighted the recency and dynamism of the field. On average, each publication received 19.2 citations, suggesting a high level of academic engagement and the relevance of the research outputs. Keyword analysis revealed a rich thematic diversity, with 23,803 keywords (plus ID) and 13,991 author keywords (DE), reflecting the breadth of topics explored. Authorship patterns demonstrated a high degree of collaboration, with 64,027 authors contributing to the corpus, including only 325 single-authored documents and 359 single-authored authors. This indicates that collaborative research dominated the field. The average number of co-authors per document was 10.3, and 24.97% of authors were involved in international collaborations, underscoring the global and interdisciplinary nature of research on long COVID. This bibliometric snapshot illustrates a rapidly expanding and highly collaborative research domain, marked by a strong focus on original contributions, significant citation impact, and a wide-ranging exploration of themes related to long COVID.

**Table 1.** Overview of long COVID research landscape from 2020 to March 2025.

| Description | Results |
| --- | --- |
| Main Information About Data |  |
| Timespan | 2020:2025 (March) |
| Documents | 9729 |
| Annual Growth Rate % | 19.81 |
| Document Average Age | 2.13 |
| Average Citations Per Doc | 19.2 |
| Document Contents |  |
| Keywords Plus (ID) | 23803 |
| Author's Keywords (DE) | 13991 |
| Authors |  |
| Authors | 64027 |
| Authors Of Single-Authored Docs | 325 |
| Authors Collaboration |  |
| Single-Authored Docs | 359 |
| Co-Authors Per Doc | 10.3 |
| International Co-Authorships % | 24.97 |
| Document Types |  |
| Article | 7867 |
| Review | 1862 |

\*DE: the frequency distribution of authors' keywords; ID: the frequency distribution of keywords associated with the manuscript by SCOPUS

### The trend of long COVID research from 2020 to March 2025

Figure 1 illustrates the trajectory of scholarly interest in long COVID over a five-year period, from 2020 to March 2025. The data revealed a consistent upward trend in publication volume from the onset of the pandemic, culminating in a peak in 2024. This surge likely reflected heightened global attention and substantial research funding directed toward understanding the long-term consequences of COVID-19. The peak in 2024 might also correspond with the maturation of early research initiatives and the publication of large-scale studies. Following this peak, the publication trend appeared to plateau, suggesting a potential shift in research priorities, a reallocation of funding, or a perception that foundational questions have been sufficiently addressed. The coefficient of determination (R² = 0.86) indicated a strong correlation between time and publication output, affirming the robustness of the observed trend. However, the apparent decline in 2025 should be interpreted with caution, as the data only encompass the first quarter of the year. It remains unclear whether this represents a temporary fluctuation or the onset of a broader reorientation in scientific focus. The figure captures the dynamic evolution of long COVID research and raises essential questions about the sustainability of academic interest in the topic. The observed trends show the need for continued monitoring to determine whether the field is entering a phase of consolidation or transitioning toward new research frontiers.

**Figure 1:**
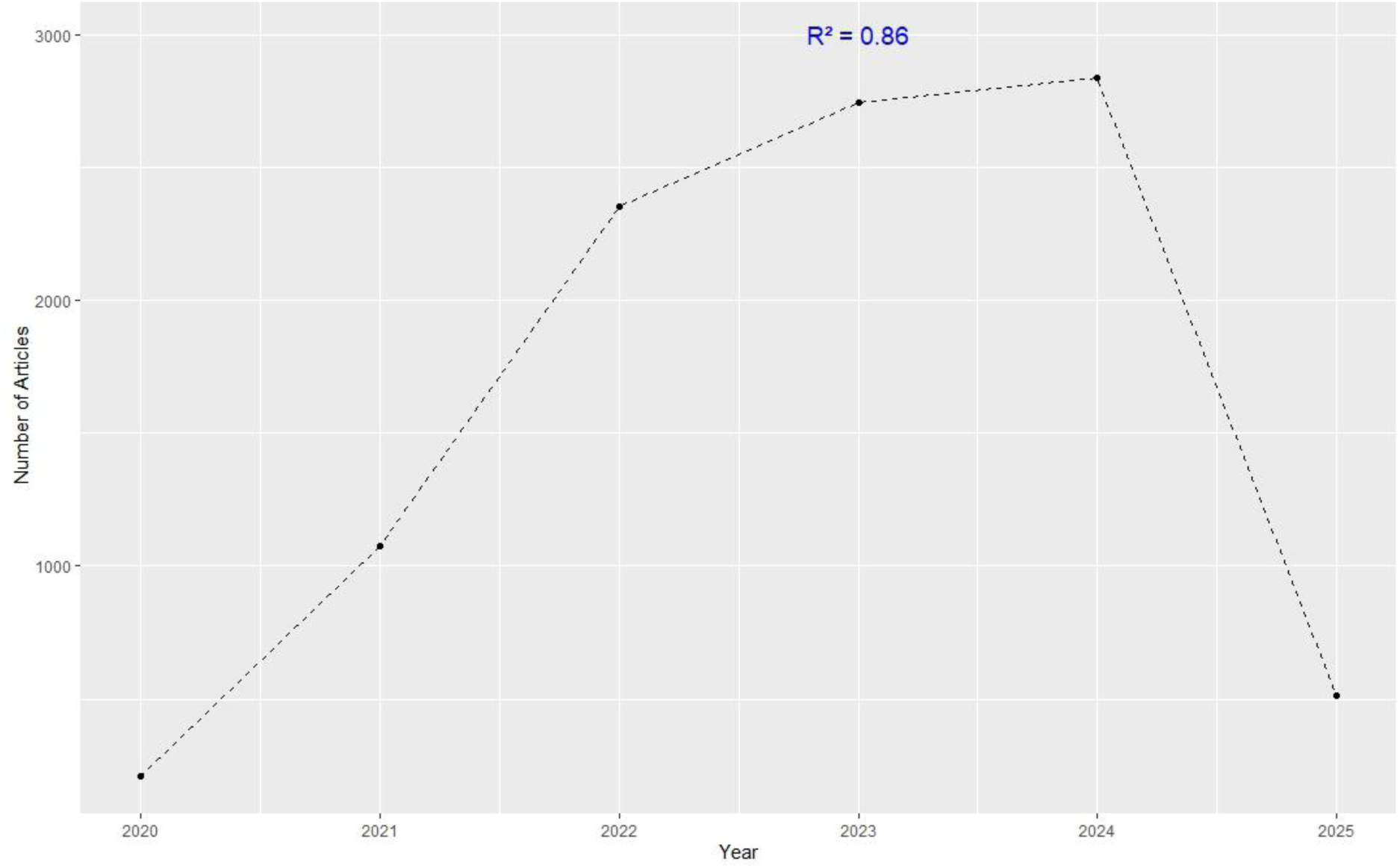
The trend of the long COVID research landscape from 2020 to March 2025

### Distribution of long COVID research by countries

Figure 2 shows that the United States had the highest number of long COVID research, China was second, with the United Kingdom and Italy rounding out the following positions. Other countries, such as India, Spain, and Turkey, made smaller yet significant contributions, while nations like Germany, Iran, and Brazil were at the lower end of the chart. From Figure 2, a few assumptions can be made: i) The dominance of the United States and China could reflect their strong research ecosystems, funding availability, and large academic communities; and ii) countries with lower contributions, like Iran and Brazil, might indicate areas where international collaboration or additional resources could help increase research output.

**Figure 2:**
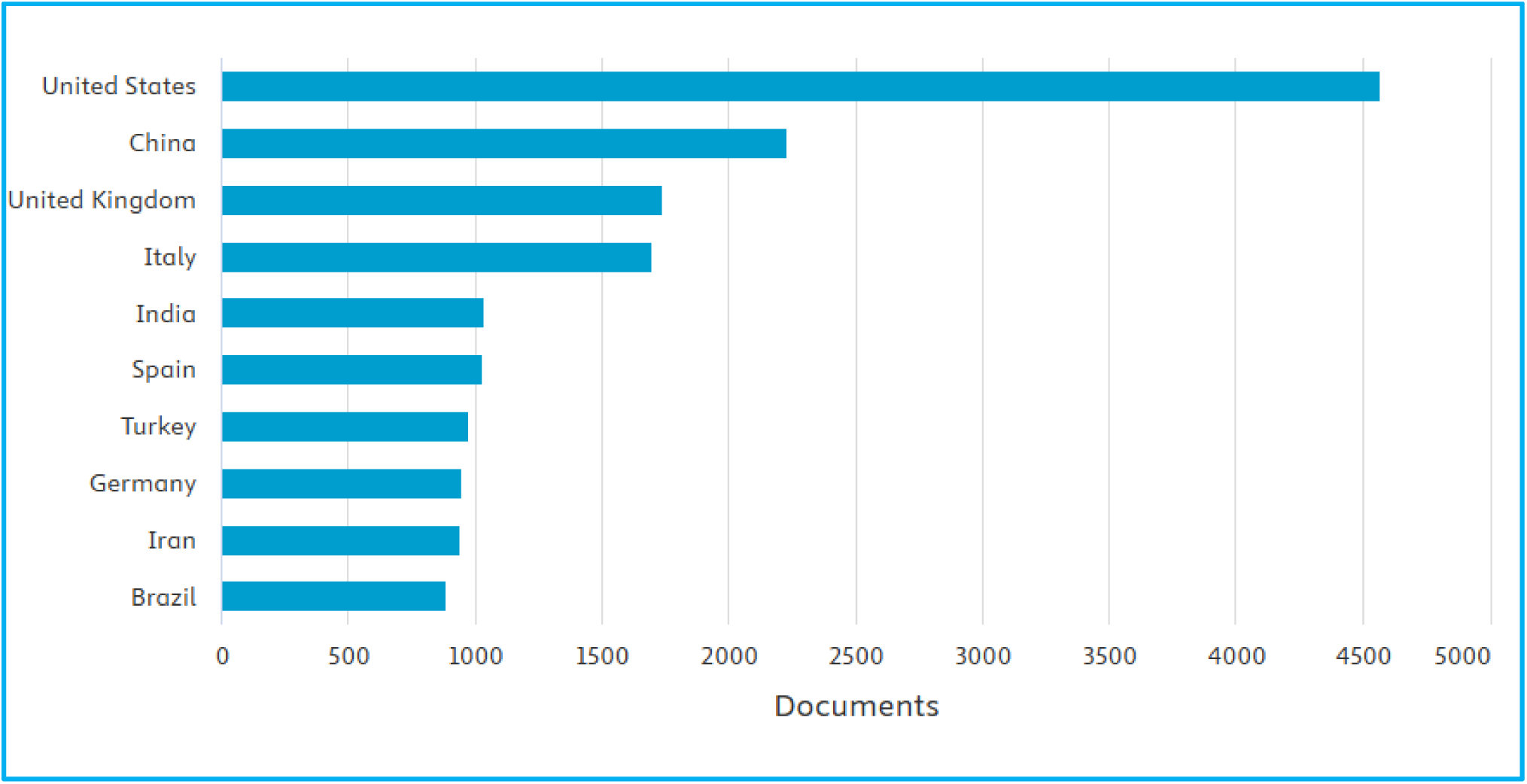
Distribution of long COVID research by the top 10 countries.

### Influential authors, institutions, and journals

Figure 3 provided a visual representation of the contributions made by different authors in terms of the number of documents they had produced. Wang Y had the highest number of publications,82, making the author the most relevant author in long COVID research. Zhang Y, with 72 publications, followed closely behind Li Y, with 60, and Li X, with 59. The remaining authors displayed a range of contributions, creating a gradient of productivity across the field. This plot is handy for highlighting the dominance or influence of specific individuals in a particular area of research. The plot allows for quick comparison and sheds light on potential key figures driving the discourse or advancing the field.

**Figure 3:**
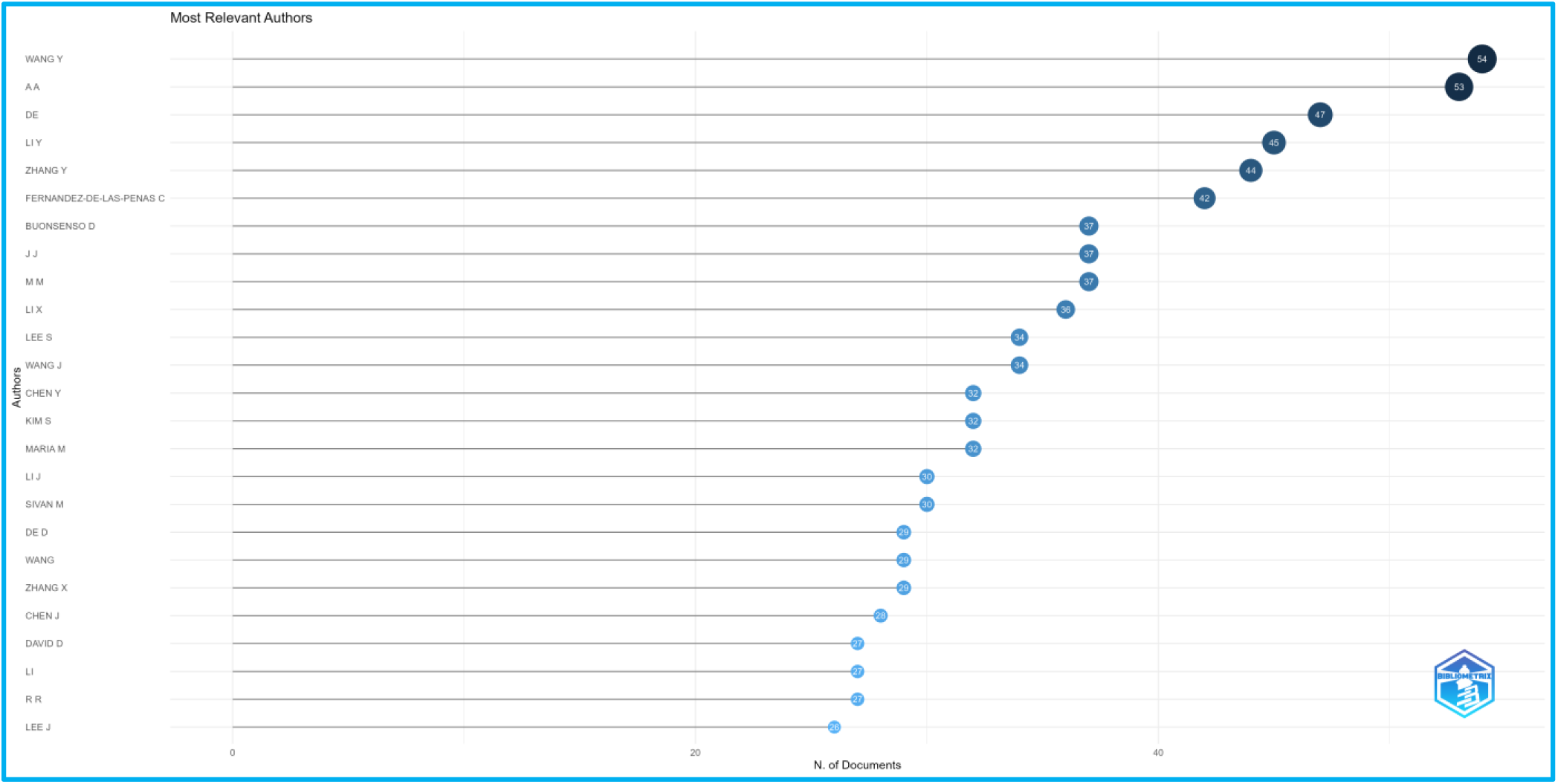
Most relevant authors in long COVID research

Figure 4 revealed the significant contributions of leading academic and research institutions in long COVID research from 2020 to 2025. The University of California had topped the list, with 577 publications showcasing its pivotal role in advancing understanding and solutions for this complex condition. Oxford University (498 articles) and the Icahn School of Medicine at Mount Sinai (464 articles) followed closely, indicating substantial research output from these leading global healthcare and medical institutions. University College London (452), Harvard Medical School (442), University of Toronto (353), and Sapienza University of Rome (329) all made significant contributions. The presence of institutions like the Mayo Clinic (306), University of Leicester (293), and University of Milan (285) underlined the diversity of contributors spanning different regions and specialisations. This distribution of publications illustrates a concentrated effort in long COVID research by these institutions, likely driven by their access to resources, expertise, and opportunities for collaboration. It also raises questions about how this research output is being translated into treatments or policies to combat the long-term effects of COVID-19.

**Figure 4:**
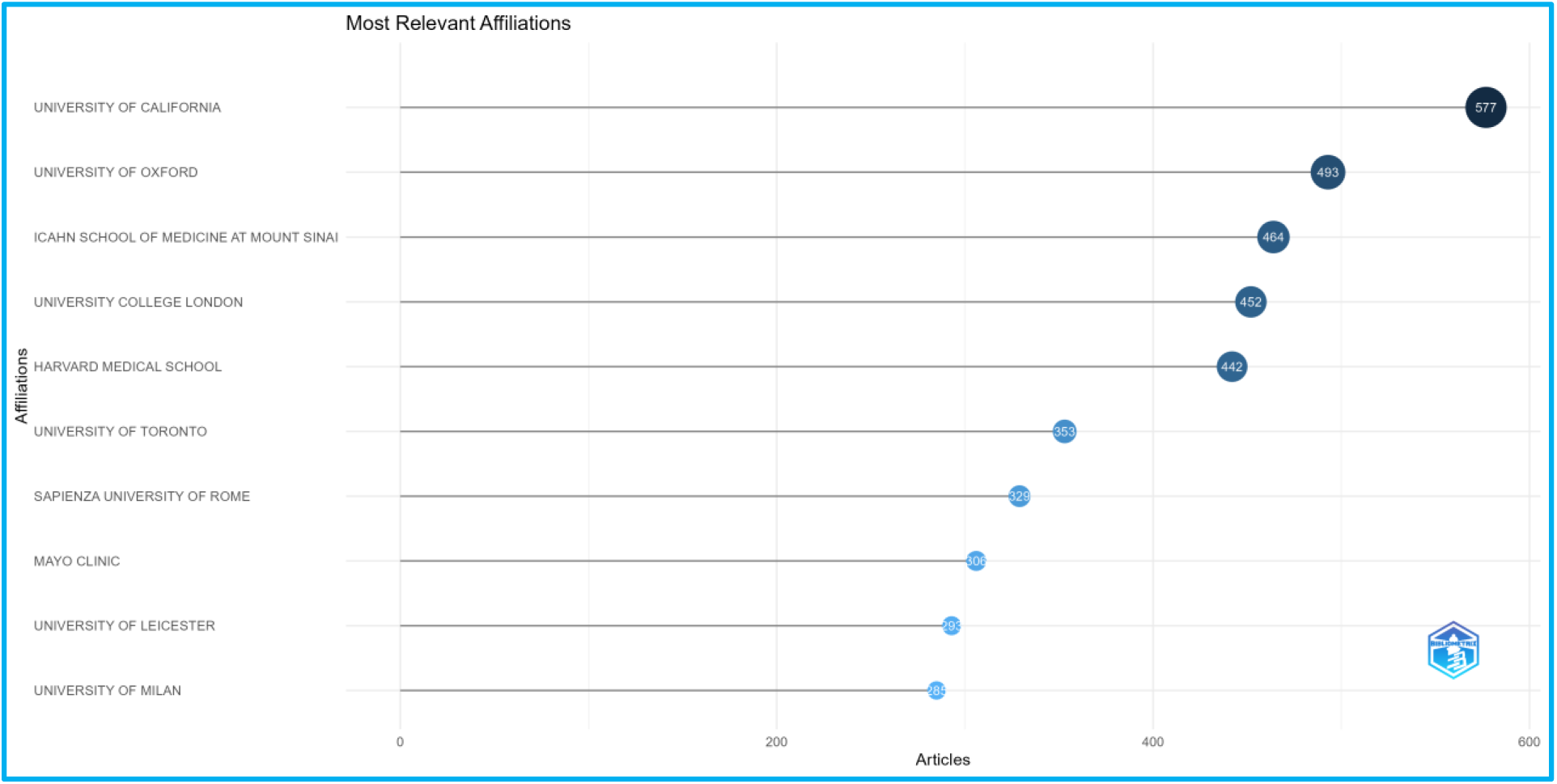
The most relevant institutions in long COVID research

### Co-word analysis and keyword clustering

This network visualisation of keyword co-occurrences provided an insightful overview of the themes and interconnections within long COVID research from 2020 to 2025 (Figure 5). The size of each keyword reflects its frequency, showing the prominence of specific topics. At the same time, the connecting lines indicate their co-occurrence in the research literature, emphasising the interconnected nature of these themes. The blue cluster focuses on broad concepts such as “pandemic,” “mental health,” and “epidemiology,” highlighting research into the general and societal impacts of long COVID. The green cluster delves into clinical aspects, featuring terms such as “inflammation,” “complication,” and “immune response,” which underscore the biological and physiological dimensions of the condition. The purple cluster centres on quality-of-life concerns, including “fatigue,” “depression,” and “dyspnoea,” reflecting the patient-centred challenges and symptoms associated with long COVID. This visualisation is a valuable tool for identifying key research areas and trends. It revealed how diverse fields, from mental health to immunology, were interconnected in studying long COVID, encouraging multidisciplinary collaboration. The network visualisation also sheds light on potential research gaps or underexplored areas, guiding future investigations.

**Figure 5:**
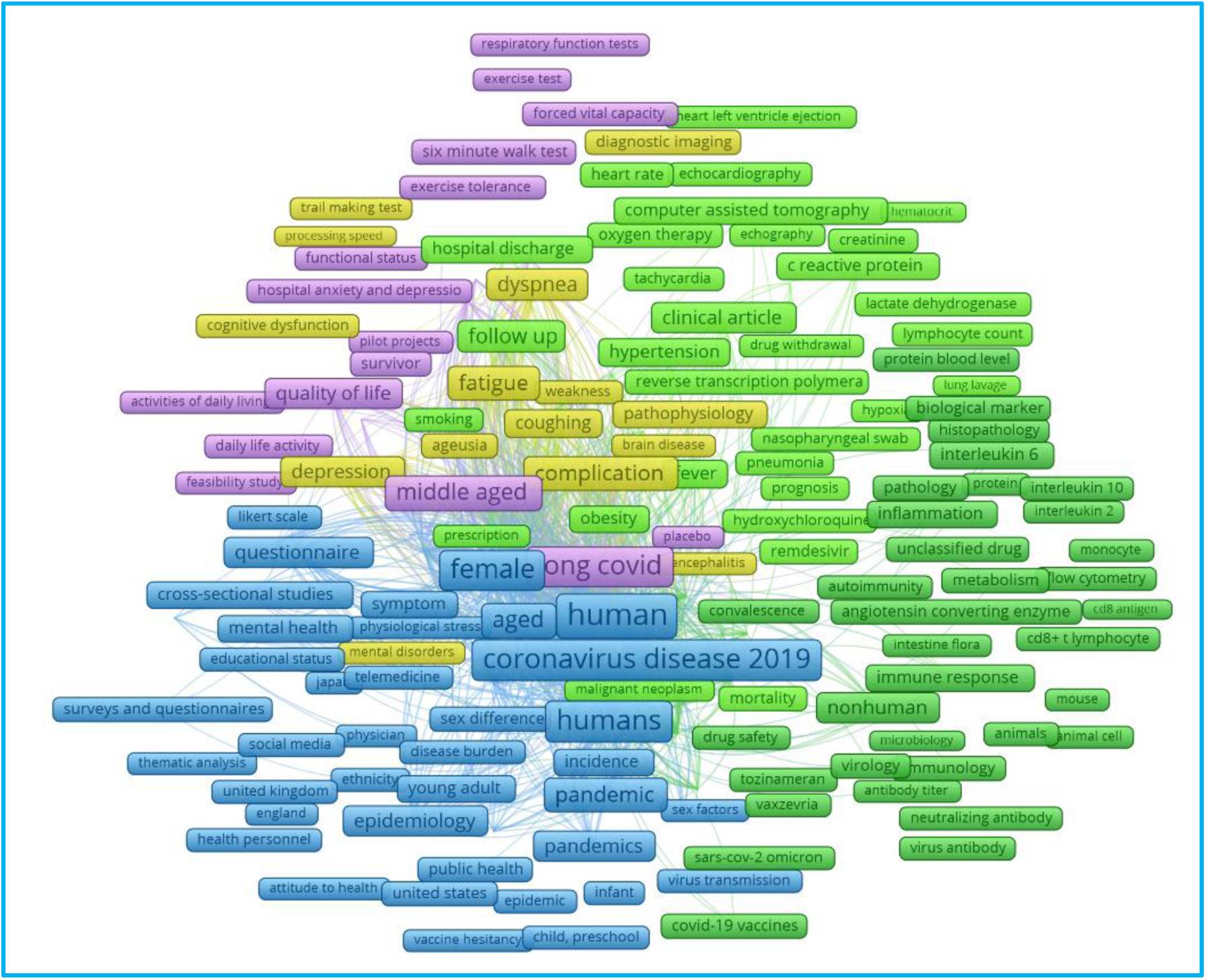
Network visualisation of keywords in long COVID research

### Citation and co-authorship analysis

The central node, “Fernández-de-las-Peñas, César,” was a significant contributor, indicating high collaboration and influence in long COVID research (Figure 6). Surrounding clusters, differentiated by colour, represented groups of authors with interconnected works, suggesting thematic research communities or shared research interests. The prominence of the clusters revealed the importance of interdisciplinary approaches in long COVID research. Larger nodes likely correlated to more publications or greater impact within the research network, while smaller nodes and isolated connections might indicate niche topics or emerging contributors. The citation by authors visualisation provided insights into how knowledge was disseminated across the research community, revealing hubs of influence and collaboration that could drive innovation and exploration. It could serve as a roadmap for researchers looking to identify key collaborators or unexplored areas within the study of long COVID.

**Figure 6:**
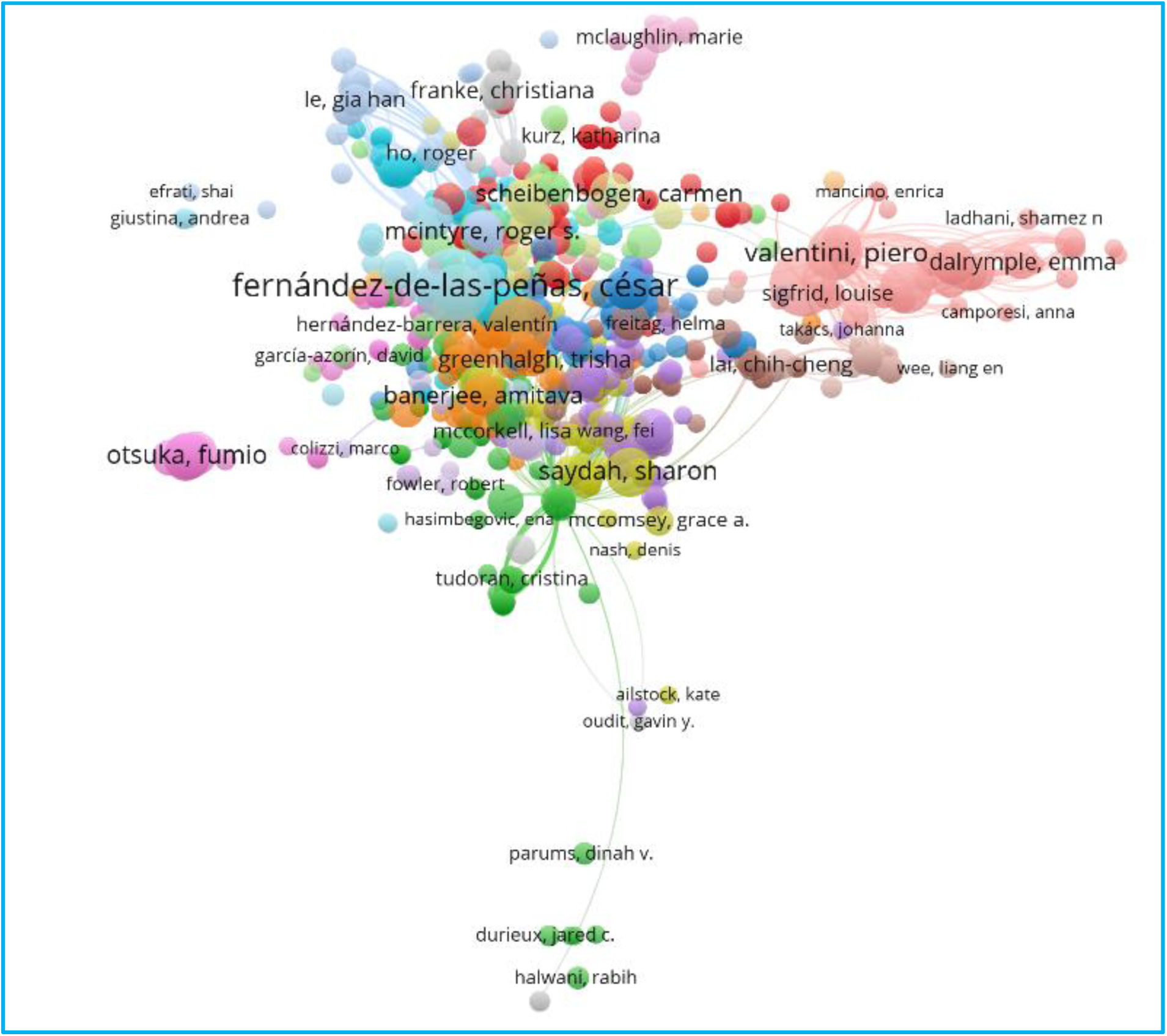
Citation by authors. The visualisation highlights collaborative research dynamics in long COVID studies from 2020 to 2025.

Journals such as Frontiers in Immunology, Journal of Clinical Medicine, International Journal of Environmental Research and PLOS ONE appeared to be central players, with larger nodes indicating their prominence and wide recognition in the research community, as shown in Figure 7. The presence of colour-coded clusters signifies distinct yet interrelated scientific fields. The interconnectedness of the clusters highlights the interdisciplinary nature of this research domain. Studies on long COVID transcend traditional boundaries, combining fields like neurology, paediatrics, immunology, and public health to provide a holistic understanding. This visualisation further highlights potential research pathways: pinpointing key journals for publishing and information sourcing.

**Figure 7:**
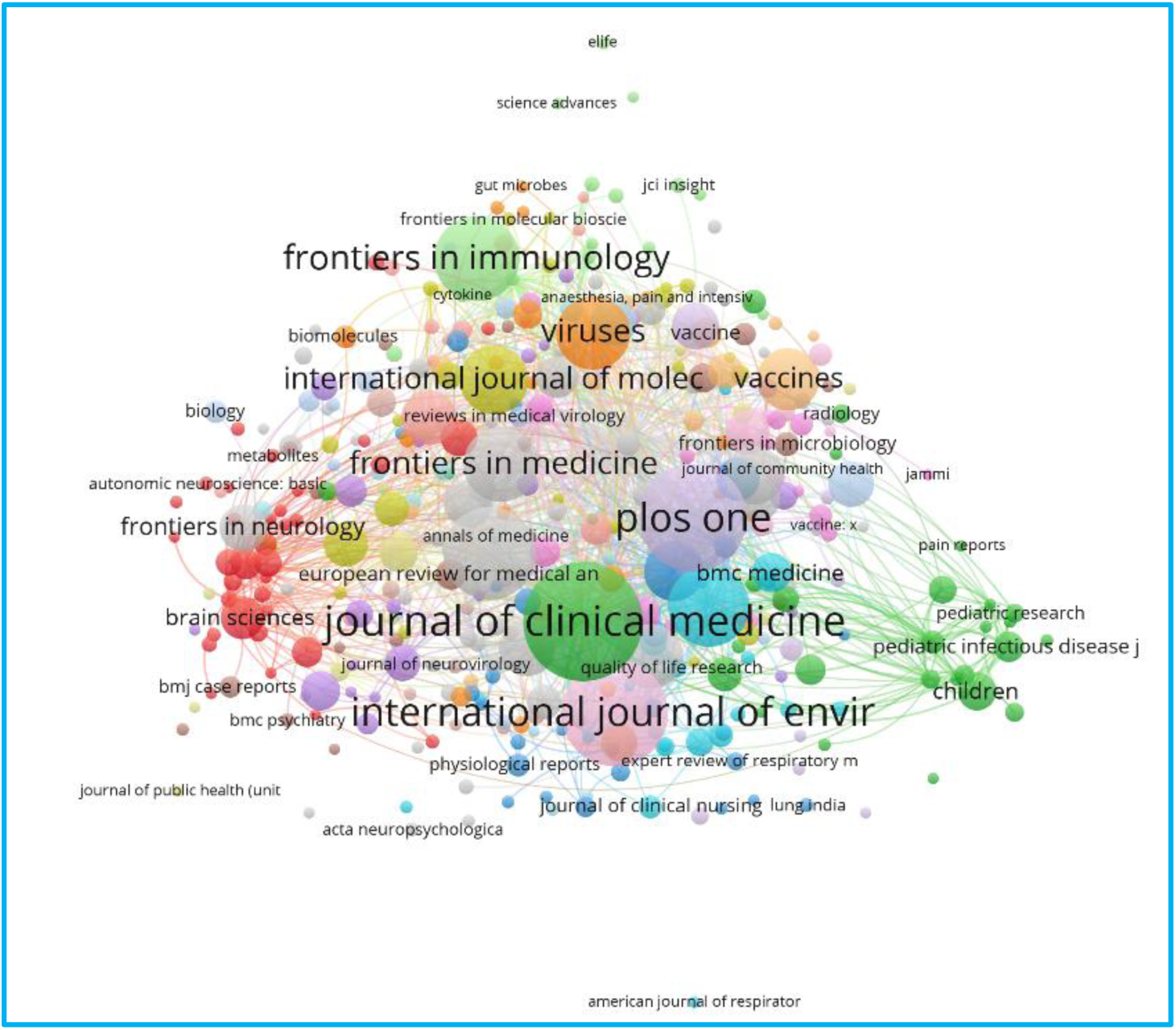
Citation by sources captures the web of co-citations among scientific journals from 2020 to 2025.

Each node represents an author, as shown in Figure 8, with the node’s size reflecting their prominence based on the number of publications or citations they have. Larger nodes, such as “Davis (2023),” “Nalbandian (2021),” “Xie (2022b),” and “Yong (2021b),” were major contributors to the research on long COVID. The clusters were colour-coded, suggesting groups of authors who frequently collaborated or whose work was closely related. This visualisation highlights the multidisciplinary nature of long COVID research, with diverse experts collaborating to tackle complex challenges. It also identified potential opportunities for new researchers to collaborate by showcasing underexplored connections or isolated nodes.

**Figure 8.**
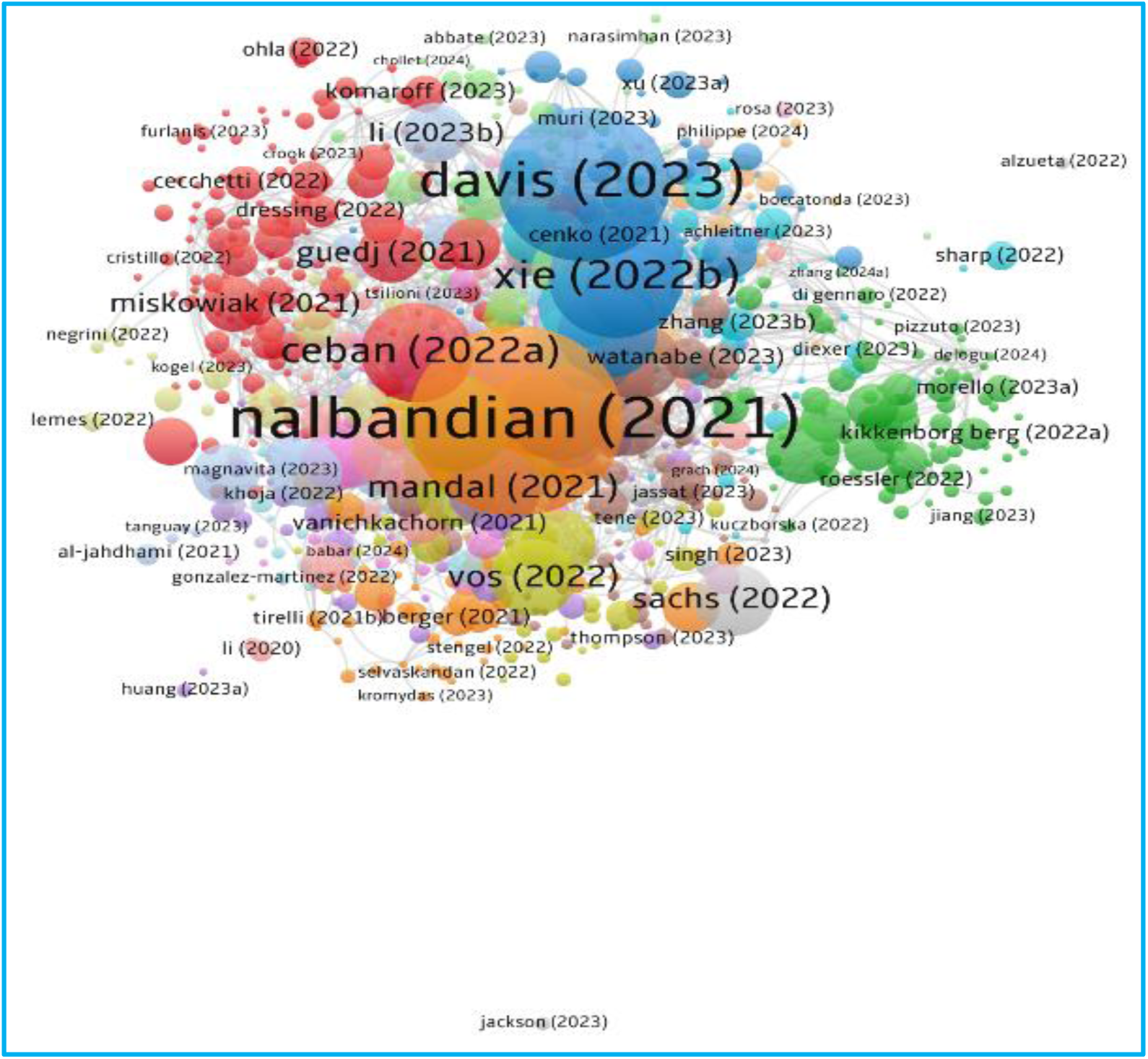
Citation by document: This visualisation highlights the network of influential authors contributing to long COVID research, offering insights into collaboration patterns and the relative impact of key researchers.

### Citation by organisation

Each node represented an academic or research institution, while the lines connecting them indicated collaborative relationships (Figure 9). The size of the nodes reflected the prominence or influence of these institutions within the research network, with larger nodes signifying a greater contribution or recognition in long COVID studies. Notable entities like Harvard Medical School, Mayo Clinic, and University College London appeared as central hubs, suggesting they were key players in this research domain. Their positions and connections highlight their role as centres of collaboration and knowledge dissemination. The colour-coded clusters point to thematic collaborations or specialities among institutions. The presence of diverse institutions from various countries underscores the global nature of long COVID research. This visualisation emphasises the importance of institutional collaboration in advancing long COVID research. It also reveals how diverse academic and clinical centres pool their expertise, bridging gaps between fields like immunology, public health, and clinical medicine. In addition, it helps identify influential organisations or emerging contributors that could drive future research efforts.

**Figure 9:**
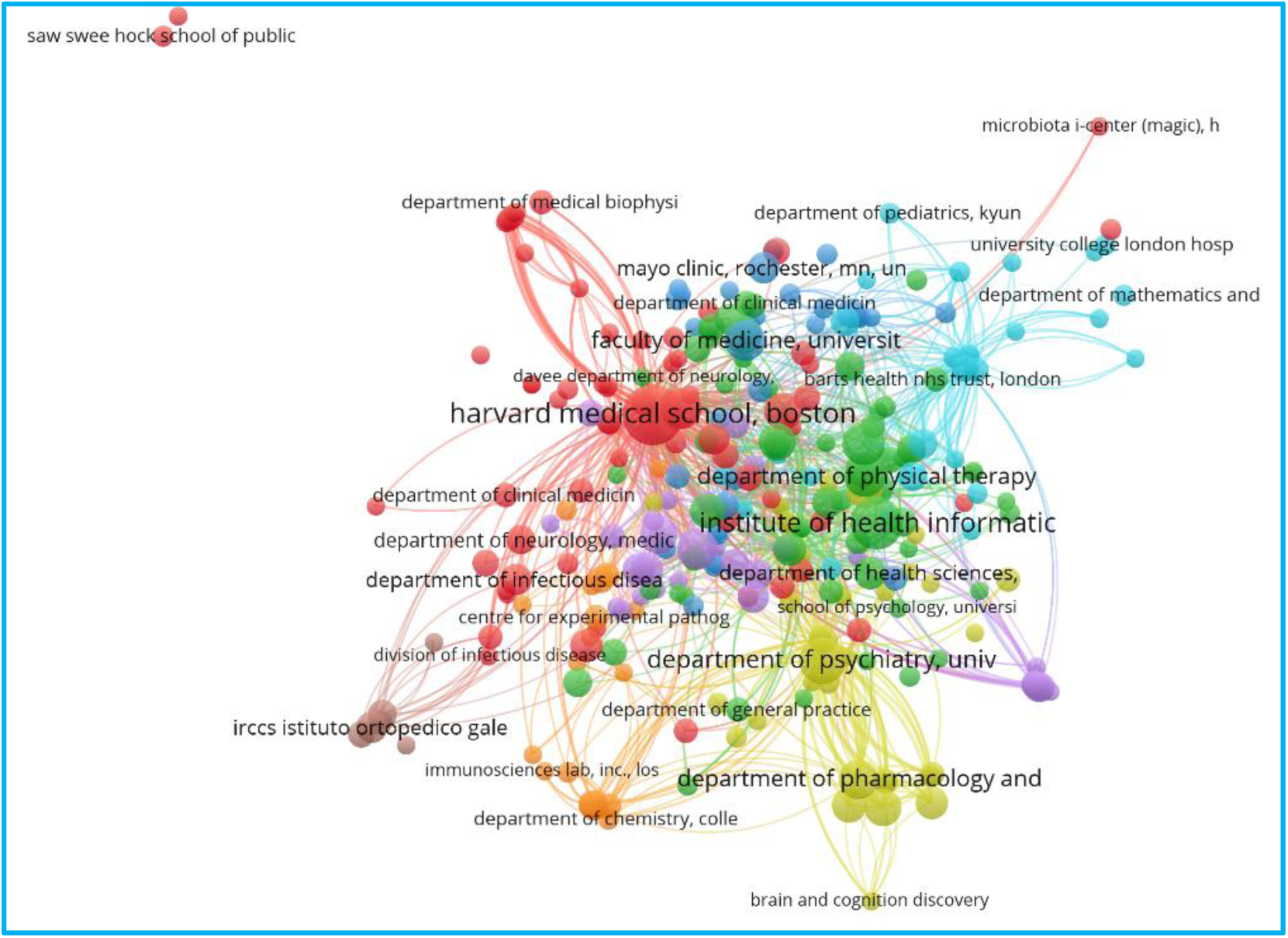
Citation by organisation (academic and research institutions)

### Citation by countries

In Figure 10, the United States was the most prominent node, suggesting its central role in fostering international collaborations. Its extensive connections with countries like the United Kingdom, Germany, and China highlighted its influence and involvement in global research. Countries like the United Kingdom, Germany, France, Italy, and Spain formed a cohesive group, indicating strong regional partnerships in Europe. Smaller nodes represented countries with less frequent collaborations or emerging research activities. This highlights areas where efforts could be intensified to involve a broader range of nations. This visual emphasises the interdisciplinary nature of solving complex worldwide problems. Examples of bridging knowledge gaps and sharing resources include collaborations between developed and developing countries.

**Figure 10:**
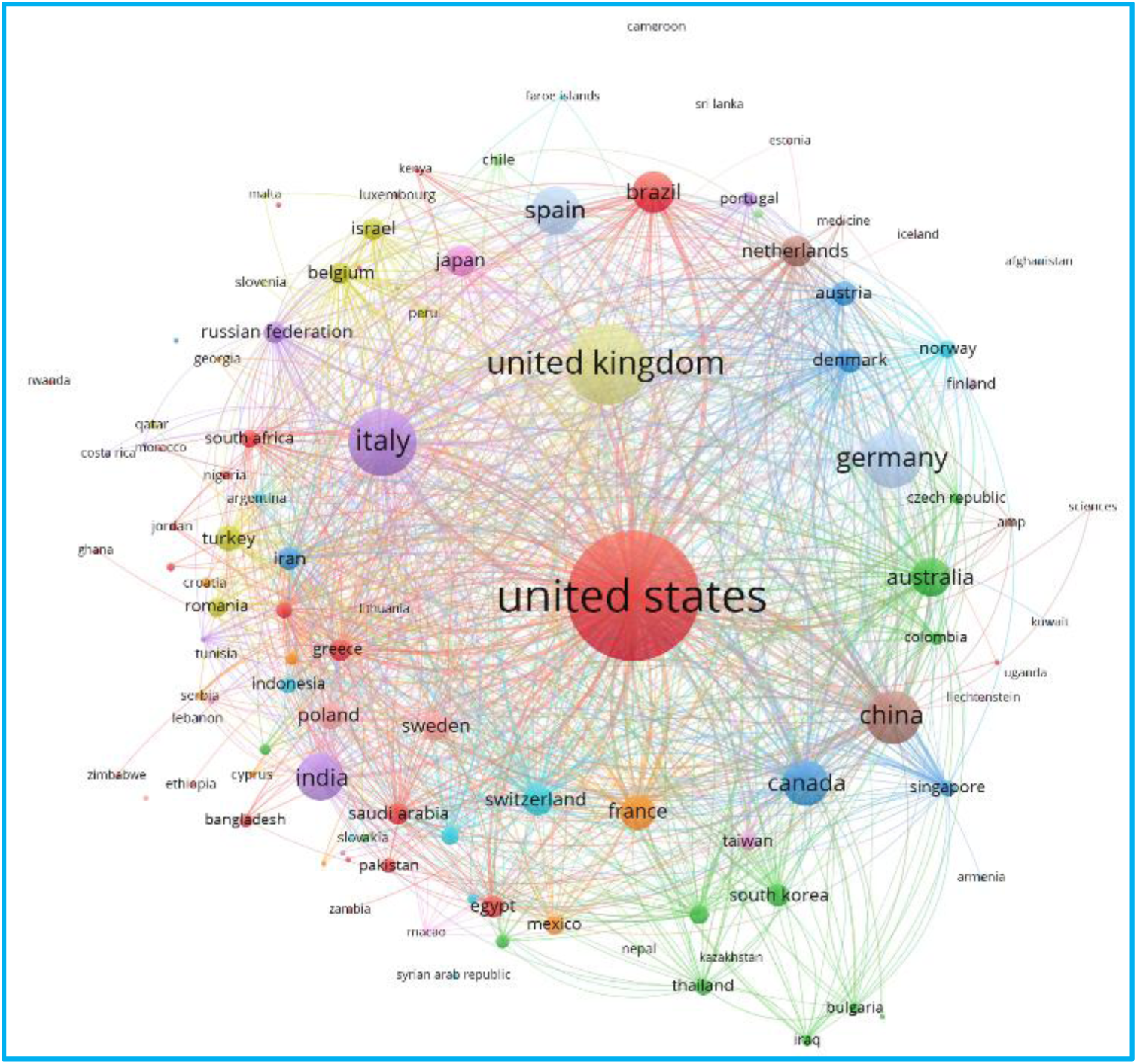
Citation by Countries. Maps the global collaboration network in long COVID research, as inferred from the connections between countries. It maps the global collaboration network in long COVID research, as inferred from the connections between researchers.

### Co-authorship

#### Co-authorship by authors

Larger nodes represented authors with significant contributions through numerous publications, as shown in Figure 11. These authors acted as central figures in the research network. Smaller nodes indicated less frequent collaboration or emerging contributors specialising in niche areas. The colour-coded clusters reflected distinct research groups or thematic communities. This segmentation revealed the interdisciplinary approach to understanding and addressing the condition. The links between nodes indicate co-authorship ties, suggesting that these authors frequently collaborate on shared research topics. Denser connections within a cluster signify strong collaborative relationships, while sparser links could suggest occasional partnerships. This visualisation revealed the significance of collaboration in scientific advancement. Key researchers served as hubs, fostering innovation and linking diverse thematic areas.

**Figure 11:**
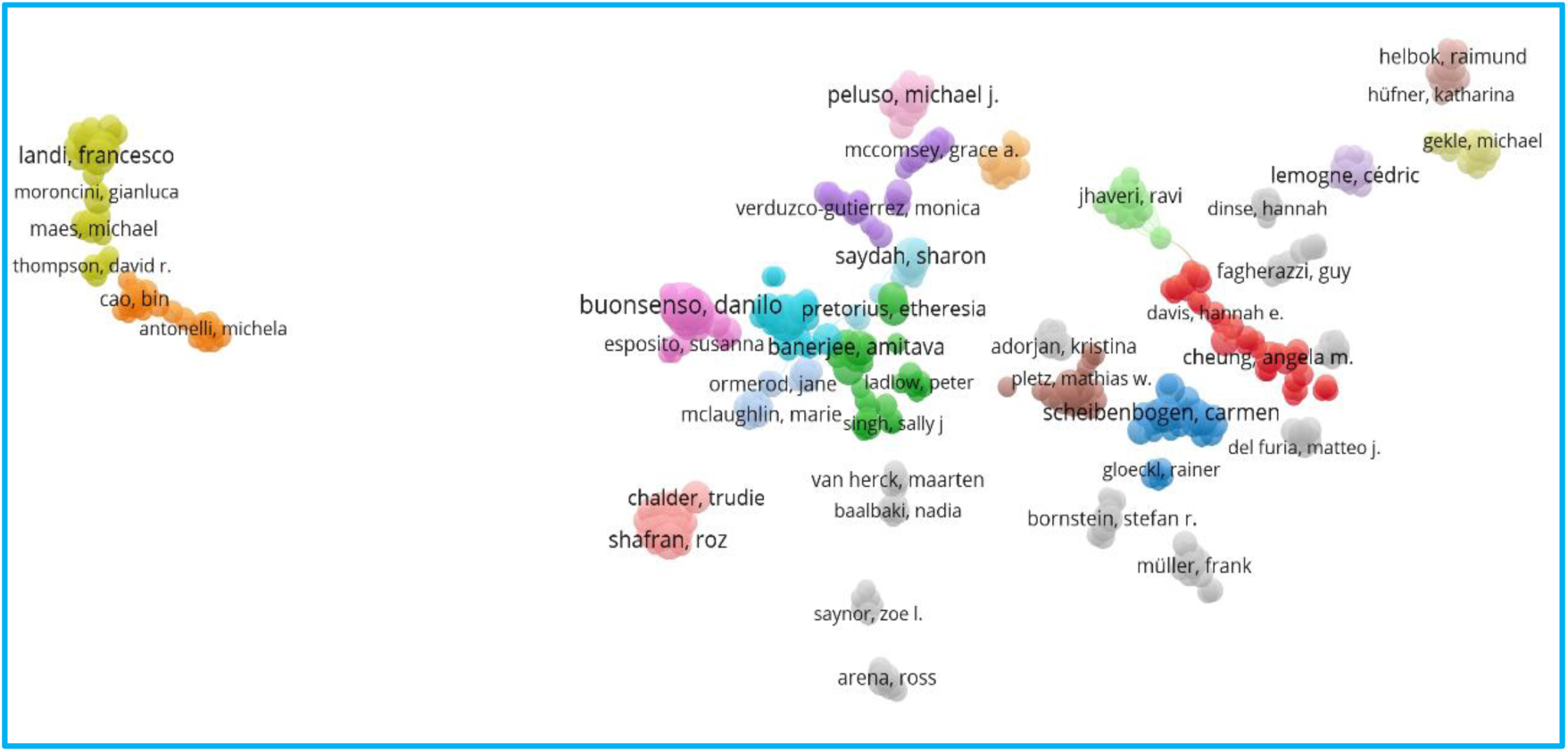
Co-authorship relationships among researchers, emphasising the collaborative dynamics within long COVID studies.

#### Co-authorship by countries

The United States was a central hub, with extensive connections to nations such as China, Italy, Canada, and Australia, as shown in Figure 12. This suggests the U.S. is pivotal in funding, leading, or collaborating internationally. European nations, such as the United Kingdom, Germany, and France, formed a well-connected cluster, indicating robust regional partnerships. Emerging countries, represented by smaller nodes, have shown growing participation in this research, which signifies an expanding interest and capacity.

**Figure 12:**
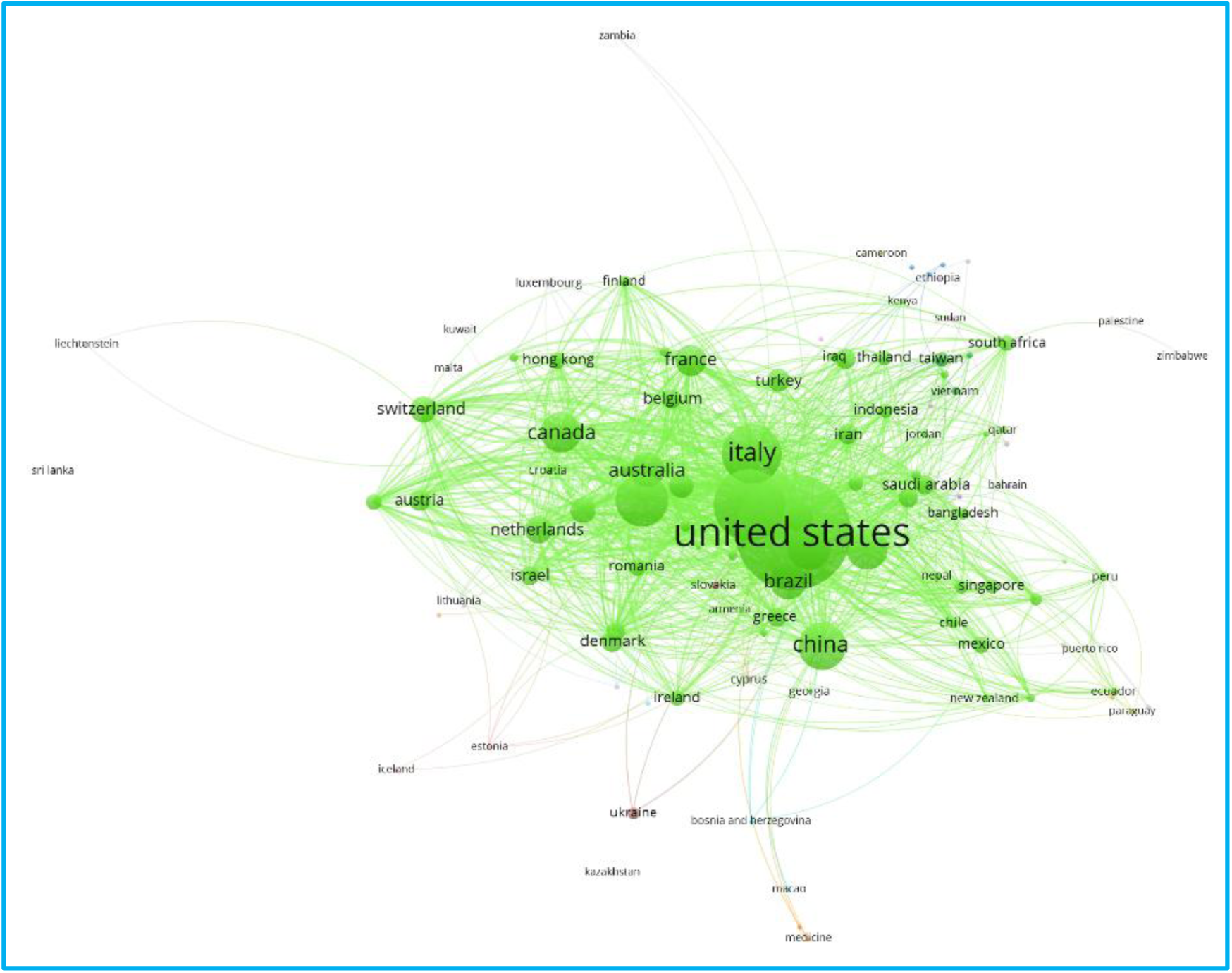
Co-authorship by countries

### Implications of the findings for clinical practices, policy and future research

The findings of this bibliometric analysis carry several important clinical implications. The significant annual growth rate of long COVID publications (19.81%), peaking in 2024, reflects the heightened global urgency to understand and manage this condition. For clinicians, this expanding evidence base shows the importance of staying current with rapidly evolving research to provide informed, evidence-based care. It also suggests that clinical guidelines and best practices will continue to adapt as new knowledge emerges, requiring flexibility in clinical decision-making. These findings also reflect the urgency with which governments and health systems should prioritise this condition. Policymakers should recognise long COVID as a long-term public health challenge that requires sustained investment in research, surveillance, and health services delivery. The expansion of evidence-based provides an opportunity for governments to integrate emerging knowledge into national health strategies, ensuring that guidelines remain up to date and responsive to new insights.

The identification of the United States, China, and the United Kingdom as leading contributors to long COVID research highlights both opportunities and challenges for clinical practice. While these countries are generating valuable insights, much of the evidence is derived from high-income settings and may not fully capture the clinical realities in low- and middle-income countries. Clinicians should therefore interpret available evidence within the context of local resources, patient populations, and health system constraints, while advocating for more inclusive and diverse research collaborations. Policymakers in low- and middle-income countries (LMICs) may risk adopting recommendations derived from contexts with very different health system capacities, patient demographics, and socio-economic realities due to the uneven global distribution of scientific activity. This finding highlights the need for targeted funding and capacity-building initiatives to foster inclusive international collaborations that generate contextually relevant evidence for diverse populations. Strengthening global partnerships and fostering inclusive research networks will be essential for equitable knowledge production.

The wide thematic range of studies, spanning clinical, psychological, and social aspects of long COVID, reinforces the complex and heterogeneous nature of the condition. This has direct implications for patient care, as it necessitates interdisciplinary approaches that integrate medical treatment with psychological support and social care. Multidisciplinary clinics and coordinated care pathways are particularly well-suited to address the heterogeneous needs of patients with long COVID. Therefore, policymakers should move beyond biomedical frameworks to design integrated care pathways that incorporate mental health services, social support, rehabilitation, and workplace policies. This approach aligns with the broader principle of universal health coverage, ensuring that patients’ complex needs are adequately addressed across health and social care systems. Longitudinal and patient-centred studies are needed to deepen understanding and guide clinical care. The field also benefits from strong interdisciplinary engagement, drawing on medicine, public health, neuroscience, and social sciences. Continued integration of diverse expertise will be key to addressing the multifaceted nature of long COVID.

The role of key institutions, such as the University of California and Oxford University, in advancing long COVID research also signals where landmark trials, treatment guidelines, and consensus statements are likely to originate. Clinicians should remain attentive to outputs from such research hubs to align practice with emerging evidence. The prominence of these leading institutions in driving the research agenda further points to the importance of institutional support and strategic investment in research infrastructure. National governments and funding agencies should strengthen research capacity within their own contexts, encouraging knowledge exchange and supporting early-career researchers in long COVID and related fields. The gaps identified in the literature, particularly in underexplored populations, long-term follow-up, and intervention effectiveness, signal areas where policy direction is needed. Policymakers can use such evidence gaps to shape funding priorities, support national registries, and incentivise clinical trials that evaluate therapeutic and rehabilitative strategies. With attention to these gaps, health policy can ensure that responses to long COVID are not only evidence-based but also equitable, sustainable, and globally relevant.

### Limitations of the study

The study relied solely on Scopus, which, despite its breadth, may not capture all relevant literature, particularly from regional journals, non-English sources, and open-access platforms. The exclusion of grey literature, such as preprints, reports, and conference proceedings, also limits the scope of the analysis and may omit emerging or informal contributions to the field. Temporal constraints are another consideration. The dataset ends in March 2025, potentially missing recent developments and trends. Moreover, publication delays, especially for high-impact studies, may distort the apparent trajectory of research output in the later stages of the study period. Language and geographic biases are evident, as the focus on English-language publications may underrepresent contributions from non-English-speaking regions. The dominance of high-income countries in publication output likely reflects disparities in research infrastructure and funding rather than a comprehensive global effort. Interdisciplinary challenges further complicate the analysis. Assigning publications to discrete disciplines may oversimplify the inherently cross-cutting nature of long COVID research, and keyword clustering may not fully capture thematic overlaps or the nuanced interconnections among research topics. These limitations underscore the need for cautious interpretation and suggest that future bibliometric studies should incorporate more inclusive data sources, account for publication lags, and adopt methods that better reflect the interdisciplinary and global character of long COVID research.

## Conclusion

In conclusion, the findings of this bibliometric analysis highlight the dynamic and collaborative nature of long COVID research and its significant impact on the scientific community. Nevertheless, they highlight the importance of sustained funding, cross-disciplinary teamwork, and concentrated efforts to fill research gaps, thus fully understanding and mitigating COVID-19’s long-term effects. By building on the strengths of existing research and addressing its limitations, future studies can contribute to improved outcomes for individuals affected by long COVID and inform global efforts to manage this complex condition.

## Data Availability

All data produced in the present study are available upon reasonable request to the authors.

